# Quantitative Assessment of Dual and Triple Energy Window Scatter Correction in Myocardial Perfusion SPECT with a 4D Phantom

**DOI:** 10.64898/2026.04.17.26351095

**Authors:** Mounia El Bab, Albert Guvenis

## Abstract

Conflicting evidence on scatter correction (SC) methods plagues quantitative myocardial perfusion SPECT (MPI), hindering standardized clinical protocols. This simulation study, utilizing the SIMIND Monte Carlo program and a highly realistic 4D XCAT phantom, systematically evaluates Dual Energy Window (DEW, with k=0.5) and Triple Energy Window (TEW) SC techniques. We uniquely investigate their performance across various photopeak window widths (2, 4, and 6 keV) and novel overlapped/non-overlapped configurations specifically for Tc 99m MPI – parameters largely unexplored in realistic cardiac models. Images were reconstructed with OSEM under uncorrected (UC), SC, and combined attenuation and scatter corrected (ACSC) conditions. Quantitative analysis focused on signal-to-noise ratio (SNR), contrast-to-noise ratio (CNR), defect contrast, and relative noise to background (RNB). Our findings consistently show ACSC’s superior performance in CNR, SNR, and defect contrast, confirming its critical role. Interestingly, SC alone reduced noise but compromised defect contrast relative to UC, highlighting a potential trade-off without attenuation correction. Crucially, this study reveals minimal influence of photopeak window width and overlap configuration on image quality, and no significant difference between DEW and TEW across most metrics. These results provide essential evidence for optimizing quantitative MPI protocols, suggesting that for Tc-99m, the choice between DEW and TEW, and specific window settings, may be less critical than ensuring robust attenuation correction.

## 1. Introduction

### 1.1 Background

Myocardial Perfusion Imaging (MPI) using Single Photon Emission Computed Tomography (SPECT) is a clinically validated and widely adopted modality for the non-invasive assessment of coronary artery disease (CAD). By visualizing regional perfusion with radiotracers like Technetium-99m MPI assists clinicians in diagnosing ischemia, evaluating myocardial viability, and guiding therapeutic decisions. However, SPECT’s diagnostic accuracy is frequently compromised by inherent physical limitations, with photon scattering being a primary source of image degradation.

Compton scattering is the dominant interaction mechanism for gamma photons in the SPECT energy range (70– 250 keV) [1]. It occurs when a photon collides with a loosely bound electron, transferring part of its energy and changing direction [1]. This results in a scattered photon with reduced energy and a trajectory that no longer points directly toward the detector [2]. While some scattered photons are absorbed by the collimator and rejected, those scattered at smaller angles can pass through the collimator holes and be misclassified as primary photons [2, 3]. Due to their minimal energy loss, these misclassified photons remain within the main photopeak window, contributing to scatter-induced image degradation [2, 3].

This misclassification has two major consequences: a loss of spatial resolution due to incorrect photon origin localization, and an increase in background counts, which reduces contrast between healthy and ischemic myocardial tissue. Furthermore, conventional scintillation detectors’ limited energy resolution complicates reliable distinction between scattered and unscattered photons [2]. In clinical MPI, this is particularly problematic in areas adjacent to high-uptake organs like the liver, where scatters can introduce artifacts into the inferior myocardial wall, potentially leading to misdiagnosis [4]. To address these issues, various scatter correction (SC) techniques have been developed. Among the most commonly implemented in clinical and research settings are the Dual Energy Window (DEW) and Triple Energy Window (TEW) methods. DEW estimates scatter by scaling counts from a single lower energy window by a correction factor, assuming a linear relationship between scattered photons in the scatter window and those in the photopeak window. In contrast, TEW utilizes both lower and upper energy windows to estimate the scatter contribution within the main photopeak window.

### 1.2 Related Literature

Over the past three decades, numerous studies have compared DEW and TEW methods using both simulated and clinical data, yet a consensus on their relative superiority remains elusive. This section reviews key findings, highlighting the conflicting outcomes and methodological variations that underscore the need for further investigation.

Changizi et al. (2008) [5] examined TEW scatter correction in cardiac SPECT using Tc 99m in 80 patients, reporting an 8% increase in sensitivity and a 23% increase in specificity post-correction when compared to angiography. Conversely, Noori-Asl et al. (2013) [6] evaluated several Tc 99m SPECT SC techniques, including TEW (trapezoidal approximation) and DEW. While TEW improved contrast, it introduced significant background noise and reduced Signal-to-Noise Ratio (SNR), particularly for larger spheres. DEW, however, enhanced contrast while better preserving SNR [6]. A subsequent study by Noori-Asl et al. (2014) [7] using SIMIND simulations further showed that trapezoidal TEW decreased SNR and increased Relative Noise-to-Background Ratio (RNB) compared to uncorrected images, despite improving contrast in the NCAT phantom.

Rafati et al. (2017) [8] conducted a combined simulation (3D-NCAT phantom, GATE) and clinical study (46 patients) comparing DEW and TEW for Tc 99m MPI. Both methods improved image quality, with TEW demonstrating superior performance in enhancing contrast and SNR, and no significant difference in background noise between them [8]. Focusing on DEW, Noori-Asl and Bitarafan-Rajabi (2019) [9] found significant contrast improvement in both simulated (3D-NCAT, SIMIND) and clinical (45 patients) Tc 99m MPI datasets.

The importance of comprehensive correction models was highlighted by Hosny et al. (2020) [10], who found that full correction models, including attenuation correction (AC) and resolution recovery (RR) alongside SC with OSEM, significantly outperformed FBP. However, their study revealed trade-offs: transmural defect contrast was sometimes higher in non-SC reconstructions (e.g., OSEM-NC > OSEM-SC), suggesting potential overcorrection or noise-induced contrast loss. Similarly, SNR was higher in non-SC reconstructions, while SC-based reconstructions showed enhanced Contrast-to-Noise Ratio (CNR), particularly in apical slices. These findings suggest that while SC improves defect detectability via CNR, it may introduce additional noise or affect contrast unless integrated with AC and RR [10]. In contrast, Tantawy et al. (2020) [11] reported that DEW SC alone provided the best image resolution for Tc 99m MPI in a clinical SPECT/CT study, surpassing other corrections like AC and AC-SC. Ye et al. (1992) [12] also observed significant increases in both image resolution and defect-to-normal contrast when combining FBP with AC, RR, and DEW.

Beyond cardiac imaging, Miwa et al. (2022) [13] evaluated SC techniques for Tc 99m bone SPECT/CT. They found that DEW with a 20% sub-window width offered the most accurate quantification for bone-equivalent materials but caused slight image uniformity degradation. TEW with a 3% sub-window performed better in water phantoms but was less effective for bone-like backgrounds [13]. More recently, Alaei et al. (2023) [14] studied AC and SC impact on MPI using gated SPECT with a 4D-XCAT phantom and GATE simulations. Their results showed the highest image contrast with AC + TEW (trapezoidal approximation) and the highest SNR with AC + DEW, illustrating the complex interplay of correction methods based on the prioritized metric [14].

Interestingly, very recent results from Noori-Asl and Eghbal (2024) [15] suggest that DEW can, in some cases, slightly exceed TEW in specific metrics for Tc 99m. Their SIMIND simulations using both simple cardiac and realistic NCAT torso phantoms reported superior contrast improvement with DEW (22.48% for NCAT, 23.88% for simple cardiac) compared to TEW (19.43% for NCAT, 12.23% for simple cardiac) [15].

Despite these numerous studies, the broader literature still lacks consensus on the overall consequences and optimal settings of scatter correction. For example, Jeong et al. (2001) [16] found that while DEW improved contrast, it had no significant impact on left ventricular functional measurements. Galt et al. (1992) [17] indicated improved quantification with SC, whereas Khalil et al. (2004) [18] observed only enhanced image contrast with no significant SNR improvements. Moreover, optimal settings for the TEW method remain debated: Asmi et al. (2020) [19] concluded that a 20% main window with a 6 keV sub-window was optimal for Sm 153 imaging, while Bouzekraoui et al. (2019) [20] found a narrower 3 keV sub-window performed better for Gd 159.

### 1.3 Study Aim and Objectives

Despite the widespread use of Dual Energy Window (DEW) and Triple Energy Window (TEW) methods for scatter correction (SC) in myocardial perfusion imaging (MPI), no consensus exists regarding their relative superiority, and conflicting outcomes persist in the literature. These inconsistencies often stem from variations in imaging parameters, reconstruction algorithms (e.g., FBP vs. OSEM), correction techniques, and specific window configurations. Crucially, the interaction between SC and attenuation correction (AC), and how their integration affects quantitative image quality, remains insufficiently understood. These gaps highlight a pressing need for standardized and evidence-based SC protocols, particularly within the context of quantitative cardiac SPECT.

While DEW and TEW are established techniques, this study offers a novel and significant contribution by systematically comparing their performance across varied photopeak window widths (2, 4, and 6 keV) and distinct configurations (overlapped and non-overlapped) specifically for Tc 99m MPI. Prior research investigating such window variations has primarily focused on non-cardiac applications or employed simpler phantoms and alternative isotopes like Sm 153or Gd 159. Furthermore, the impact of overlapping versus non-overlapping window configurations within the TEW method has not been systematically explored in myocardial perfusion imaging, representing an unaddressed aspect in existing literature. This study is the first to systematically investigate these specific parameters for cardiac perfusion imaging within a robust simulation framework that leverages the anatomically and physiologically realistic 4D XCAT phantom. The comprehensive analysis includes uncorrected (UC), scatter corrected (SC), and combined attenuation plus scatter corrected (ACSC) conditions, all reconstructed using the OSEM algorithm.

Ultimately, this study aims to clarify previously inconsistent findings, uncover potential trade-offs, and contribute new evidence for optimizing correction strategies through detailed analysis of image quality metrics crucial to perfusion imaging.

## 2 Methods and Materials

### 2.1 Study Design

This study was conducted as a simulation study to evaluate the impact of various scatter correction techniques on quantitative myocardial perfusion SPECT imaging using the SIMIND (Simulating Medical Imaging Nuclear Detectors) Monte Carlo program (version 7.0.3). The simulations were based on the four-dimensional (4D) Extended Cardiac-Torso (XCAT) digital phantom, which models realistic human anatomy and physiological motions. A single cardiac cycle was represented by eight dynamic frames. To ensure statistical robustness and account for the inherent randomness in Monte Carlo simulations, each of these eight frames was simulated twice using different random seeds, effectively doubling the dataset for each condition. Projection data generated from these simulations were then reconstructed under three distinct correction scenarios: uncorrected (UC), where images were reconstructed without any scatter or attenuation correction; scatter corrected (SC), where images were reconstructed with scatter correction applied using either the DEW or one of the TEW configurations; and attenuation and scatter corrected (ACSC), where images were reconstructed with both attenuation correction (derived from the XCAT phantom data) and scatter correction applied.

### 2.2 XCAT Phantom Configuration

The 4D XCAT digital phantom, developed by Professor Paul Segars at Duke University, was used for all simulations [21]. This phantom models a standard adult male, representing the 50th percentile for height (175.23 cm) and weight, with anatomically realistic organ structures and options for simulating cardiac and respiratory motion [22]. For this study, the phantom was configured to generate attenuation maps (providing tissue-specific attenuation coefficients) and activity maps for each of the eight frames, representing different phases of a single cardiac cycle (total cycle duration: one second, time per frame: 0.125 s). The radioactivity distribution was set to mimic that of ^99*m*^Tc-Sestamibi in a typical MPI study [23]. The relative radioactivity concentrations used in the XCAT phantom for key organ groups are detailed in Table 1. Phantom output specifications included a pixel width of 0.15 cm, a slice width of 0.15 cm, and an array size of 256×256×256 voxels.

**Table 1:**
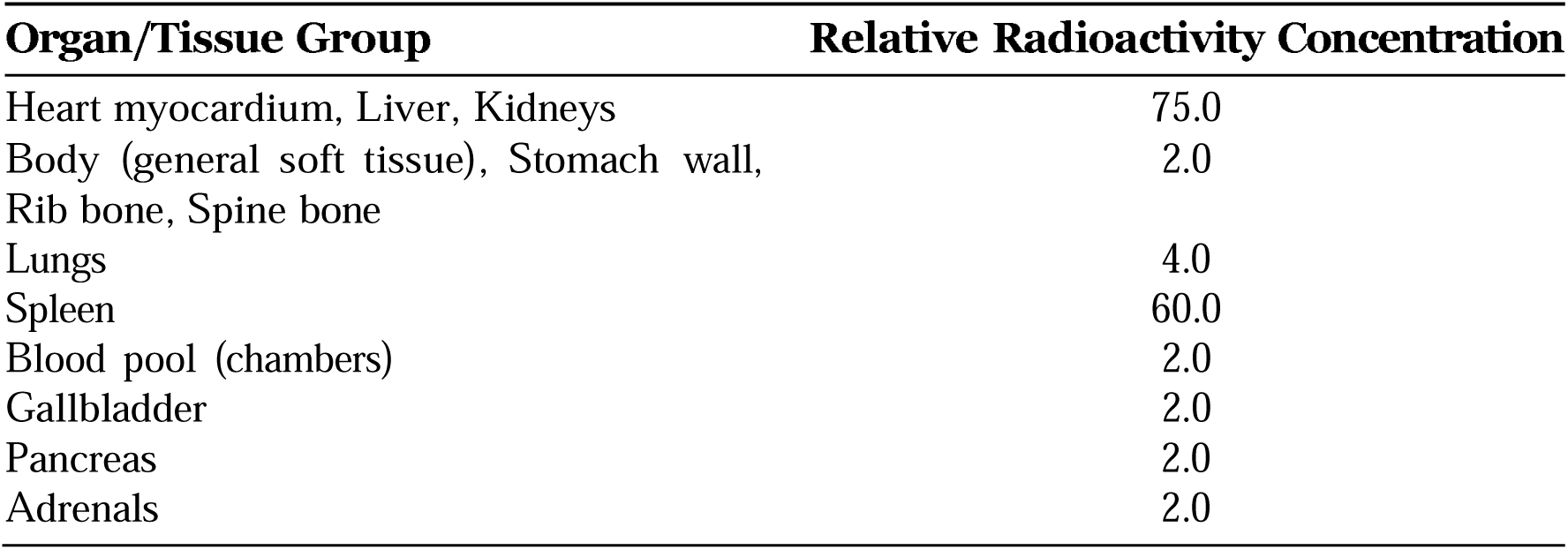
XCAT Phantom Organ Radioactivity Concentrations (Relative Units).

The anatomical modeling in this study was based on the 4D XCAT phantom, which was configured to simulate realistic cardiac dimensions and volumetric changes throughout the cardiac cycle. Table 2 presents the key morphological features of the left ventricle, including both diastolic and systolic phases. A perfusion defect was modeled in the anterolateral wall of the left ventricle (LV) to assess the impact of scatter correction on defect detectability and quantification. The lesion characteristics were adapted from a previous study having a circumferential center at 135° with a width of 140°, and a long-axis center at 0.5 (midway between base and apex) with a width of 45 mm [24]. The perfusion defect was modeled by reducing the tracer uptake in the defect region to 40% of the uptake in the surrounding myocardial tissue. Figure 1 below shows the defective phantom obtained for frame 5, illustrating the activity map for the modeled lesion in the heart. The healthy phantom configuration exhibited normal physiological left ventricular volumes: End-Diastolic Volume (EDV) of 132.1861 mL and End-Systolic Volume (ESV) of 50.5650 mL, resulting in a Stroke Volume (SV) of 81.62 mL and a Cardiac Output (CO) of approximately 4.90 L min^−1^ at a heart rate of 60 beats per minute. Detailed heart volume measurements across all frames for the healthy phantom are shown in Table 3.

**Figure 1:**
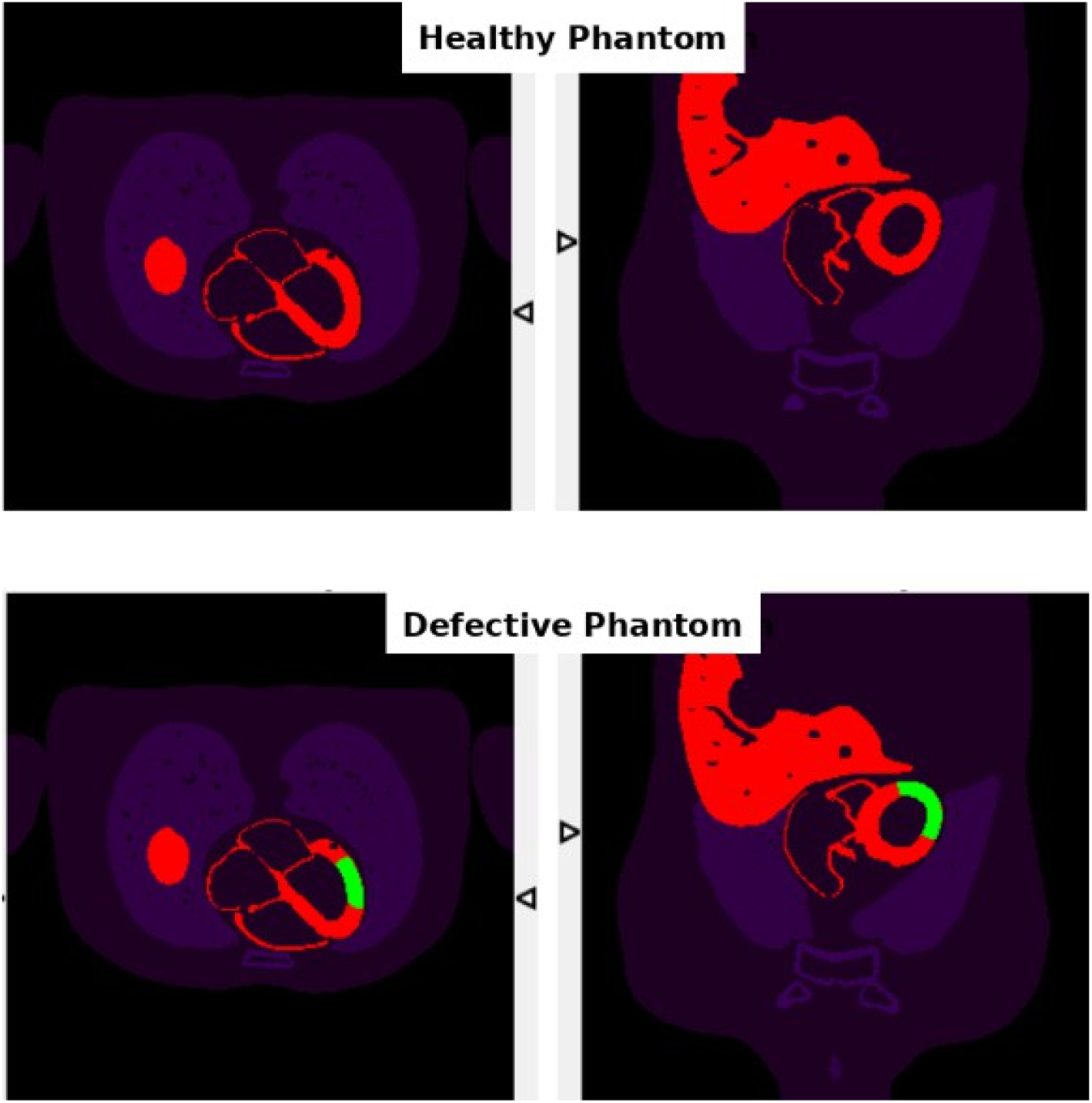
Activity map for healthy vs defective phantom for frame 5.

**Table 2:** LV Dimensions and Volumes.

| Category | Parameter | Value |
| --- | --- | --- |
| LV Size (Diastole) | Length | 96.5130 mm |
|  | Radius | 31.8726 mm |
| LV Size (Systole) | Length | 85.4235 mm |
|  | Radius | 28.6712 mm |
| LV Volumes | V1 (end-diastole) | 132.1861 mL |
|  | V2 (end-systole) | 50.5650 mL |
|  | V3 (beginning of quiet phase) | 82.7741 mL |
|  | V4 (end of quiet phase) | 109.7527 mL |
|  | V5 (reduced filling, diastole) | 123.3908 mL |

**Table 3:** Heart Volume Measurements Across Frames for Healthy Phantom.

| Frame | Time (s) | Heart Phase | Myo LV (ml) | Cham LV (ml) | Myo RV (ml) | Cham RV (ml) | Myo LA (ml) | Cham LA (ml) | Myo RA (ml) | Cham RA (ml) |
| --- | --- | --- | --- | --- | --- | --- | --- | --- | --- | --- |
| 1 | 0.000 | End-Diastole | 110.44 | 139.79 | 27.31 | 102.62 | 9.78 | 35.94 | 13.15 | 52.64 |
| 2 | 0.125 | LV 31% Contract | 119.6 | 99.3 | 25.9 | 84.44 | 11.03 | 46.63 | 14.74 | 65.99 |
| 3 | 0.250 | LV 62% Contract | 122.4 | 74.61 | 24.56 | 72.14 | 11.92 | 56.12 | 15.27 | 78.24 |
| 4 | 0.375 | LV 94% Contract | 122.58 | 59.33 | 23.33 | 63.73 | 12.57 | 63.97 | 15.69 | 88.37 |
| 5 | 0.500 | LV 17% Relax | 122.28 | 55.28 | 23.05 | 61.27 | 12.77 | 66.36 | 15.85 | 91.5 |
| 6 | 0.625 | LV 37% Relax | 122.59 | 72.82 | 24.47 | 71.28 | 11.92 | 56.92 | 15.43 | 79.05 |
| 7 | 0.750 | LV 58% Relax | 117.12 | 112.71 | 26.5 | 90.81 | 10.63 | 42.59 | 14.25 | 60.82 |
| 8 | 0.875 | LV 79% Relax | 114.31 | 125.65 | 27.01 | 96.56 | 10.25 | 39.16 | 13.75 | 56.58 |

### 2.3 Imaging Protocol for SIMIND Simulation

The study simulated a dual-head SPECT camera with specifications based on the SIEMENS Symbia T2 system [25]. A Siemens Symbia Low Energy High Resolution (LEHR) collimator with parallel hexagonal holes was modeled, coupled with a cylindrical Sodium Iodide doped with Thallium (NaI(Tl)) scintillation crystal. The simulation parameters closely followed the American Society of Nuclear Cardiology (ASNC) guidelines for a two-day stress/rest ^99m^Tc myocardial perfusion imaging protocol [26]. Key acquisition parameters are detailed in Table 4, and SPECT system specifications are in Table 5. The protocol recommends using a step-and-shoot acquisition type with a circular orbit of 180°, ranging from 45° right anterior oblique (RAO) to 45° left anterior oblique (LPO), with a minimum of 60 projections. In this study, 64 projections were simulated with a pixel size of 3 mm, which falls within the range recommended by the ASNC [26]. Additionally, the preferred matrix size according to the protocol is 128 x 128, and this was implemented in the current study. The recommended administered activity is between (296-444) MBq for a body mass index (BMI) of less than 35 kg m^−2^; therefore, the administered activity for this simulation is 370 MBq, with a projection time of 20 s for rest test [26]. The phantom CT images, which were obtained from the attenuation maps generated for the XCAT phantom, have a slice thickness of 0.15 cm and consist of 256 CT images. Projections were acquired in a 128×128 matrix with a pixel size of 0.3 cm. The system’s energy resolution was set to 9% at 140 keV, and its intrinsic spatial resolution was 0.34 cm, reflecting the typical values for contemporary Anger cameras [26]. Upon completing each simulation, an ”.smc” file was created for each frame, storing the corresponding parameters. These files will serve as inputs for image reconstruction in the subsequent step.

**Table 4:** Acquisition Parameters for SIMIND Simulation.

| Acquisition Parameter | Value |
| --- | --- |
| Orbit Type | Circular |
| Rotation Angular Range | 180° (45° RAO to 45° LPO) |
| Starting Angle | -45° |
| Rotation Angle | 2.8° |
| Radius of Rotation | 20 cm |
| Number of Projections | 64 |
| Projection Time | 20 s |
| Matrix Size | 128 x 128 |
| Pixel Size in Simulated Image | 0.3 cm |
| Intrinsic Resolution | 0.34 cm |
| Energy Window | 20% |
| Energy Resolution | 9% |
| Frames | 8 |
| Administered Activity | 370 MBq |

**Table 5:** Specifications of the Simulated SPECT System.

| Parameter | Value |
| --- | --- |
| **Collimator Parameters** |  |
| Collimator type | LEHR |
| Hole shape | Hexagonal |
| Septal thickness (mm) | 0.16 |
| Hole length (mm) | 24.05 |
| Hole diameter (mm) | 1.11 |
| Number of parallel holes | ~148,000 |
| Septal penetration (%) | 1.5 |
| **System Parameters** |  |
| System resolution @ 10 cm (mm) | 7.5 |
| System sensitivity @ 10 cm (cpm/μCi) | 202 |
| **Crystal Parameters** |  |
| Crystal material | NaI(Tl) |
| Crystal thickness (cm) | 0.9530 |
| Crystal half width (cm) | 22.25 |
| Crystal radius (cm) | 29.55 |

### 2.4 Scatter Correction Techniques

#### 2.4.1 Dual Energy Window (DEW) Method

In the Dual Energy Window (DEW) method, a single lower energy window is used to estimate the scatter contribution within the main photopeak window S*_pk_*. The estimated scatter component is calculated by scaling the counts in the lower energy window using a correction factor k, which was determined through both experimental and Monte Carlo simulation approximately to be 0.5. The value of k used in this study follows the experimental work of Jaszczak et al. (1984) [27], who demonstrated that subtracting 50% of the counts in the lower scatter window yielded compensated images with good agreement to reference data acquired in air. This value was empirically optimized for ^99m^Tc using both phantom experiments and Monte Carlo simulations on NaI(Tl)-based systems with energy resolution close to 9%, matching the specifications used in our simulation setup. Given the similarity in system geometry, isotope, and energy resolution, adopting k = 0.5 was deemed appropriate for this study. Jaszczak et al. also noted that using values of k greater than 0.5 led to overcorrection and artifact introduction, whereas k = 0.5 provided a balance between scatter removal and signal preservation [27]. Equations (1) and (2) illustrate the core principle of the DEW method, in which *T_pk_*(*i, j*) is the total number of photons detected in the photopeak window at pixel (*i, j*), and *US_pk_*(*i, j*) is the estimated count of unscattered photons [8, 6].

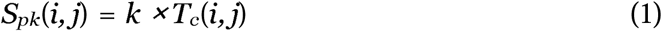

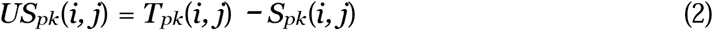

In this study, the main photopeak window was set to 126 keV to 154 keV (centered at 140 keV with a 20 percent width), while the lower scatter window ranged from 92 keV to 125 keV. For the DEW configuration, a total of sixteen simulations were performed, corresponding to eight cardiac frames, each simulated twice using different random seeds to ensure statistical robustness.

#### 2.4.2 Triple Energy Window (TEW) Method

TEW uses both lower and upper scatter windows to model the scatter distribution across the energy spectrum. The TEW method often uses a trapezoidal approximation to estimate the scatter contribution within the photopeak window. The scattered photons in the photopeak window are estimated by calculating the average counts in both the lower and higher energy windows and then normalizing them with respect to the counts in the main window [28]. In trapezoidal approximation, two narrow energy windows are used, centered at the edges of the photopeak window [7]. The estimation of the photopeak scatter spectrum involves calculating the area of a trapezoid [6]. The heights of the trapezoid’s left and right sides are calculated by the total number of photons in the upper and lower energy windows, divided by their respective window widths as shown in Equation 3, where *T_nw1_* and *T_nw2_* are the total counts in the lower and upper scatter windows, respectively, and *W_nw1_*, *W_nw2_*, *W_pk_*, represent the widths of the respective energy windows [6].

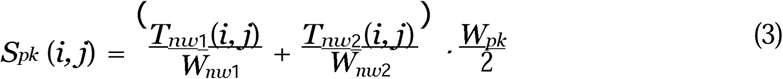

In SIMIND, this approximation is directly implemented through the “scattwin” routine, which automatically reads the defined scatter windows and calculates the scaled average of counts in the lower and upper windows, normalized by their widths and multiplied by the photopeak width, as shown in Equation 3. The lower and upper scatter windows are explicitly defined by the user in the ‘.win‘ file, which specifies the energy limits of each window. SIMIND then uses these inputs to extract the counts from the corresponding projection data and applies the trapezoidal TEW approximation accordingly. The main photopeak window remained fixed at 126 keV to 154 keV (20% width) for all TEW simulations to ensure consistent primary photon detection. Three discrete widths were tested for the TEW scatter windows: 2 keV, 4 keV, and 6 keV. The selected scatter window widths were chosen to reflect a practical range used in prior studies and to investigate their potential impact on scatter estimation accuracy. Previous work, such as by Asmi et al. [19] and Bouzekraoui et al. [20], demonstrated that sub-window widths between 2–6 keV are commonly applied in clinical or simulation-based SPECT studies, though the optimal width may vary by isotope or setup. In this study, we aimed to evaluate whether varying window widths within this commonly used range would affect image quality in the context of ^99m^Tc myocardial perfusion imaging, particularly when combined with attenuation correction.

Each configuration included both non-overlapped and overlapped versions for scatter windows of 2, 4, and 6 keV widths. In the non-overlapped setup, the scatter windows were placed immediately adjacent to the main photopeak window (126–154 keV), ensuring no intersection. In the overlapped configuration, the scatter windows were shifted inward such that each one extended 1–3 keV into the photopeak window, depending on window width. For example, with a 2 keV window, the overlapped lower scatter window spanned 125–127 keV (1 keV overlap), while the non-overlapped version was 124–126 keV (no overlap). This resulted in six unique TEW configurations per frame (3 widths x 2 overlap conditions). With eight dynamic frames, each simulated twice, a total of ninety-six TEW-based simulations were performed (6 configurations x 8 frames x 2 runs). Table 6 summarizes the TEW simulation window settings.

**Table 6:** Summary of TEW Simulation Window Configurations (Main Photopeak: 126 keV to 154 keV).

| Scatter Window Width | Overlap Type | Lower Window (keV) | Upper Window (keV) |
| --- | --- | --- | --- |
| 2 keV | Non-Overlapped | 124–126 | 154–156 |
|  | Overlapped | 125–127 | 153–155 |
| 4 keV | Non-Overlapped | 122–126 | 154–158 |
|  | Overlapped | 124–128 | 152–156 |
| 6 keV | Non-Overlapped | 120–126 | 154–160 |
|  | Overlapped | 123–129 | 151–157 |

### 2.5 Image Reconstruction

Raw projection data from SIMIND (stored as ”.smc” files) were first converted into a format compatible with the CASTOR (Customizable and Advanced Software for Tomographic Reconstruction) software using the ’Smc2castor’ utility provided within the SIMIND package [29]. A total of 112 unique simulation datasets (16 DEW + 96 TEW) were processed. Each dataset was reconstructed using the Ordered Subsets Expectation Maximization (OSEM) algorithm implemented in CASTOR. Based on prior research suggesting optimal diagnostic accuracy with such configurations for myocardial perfusion imaging, a consistent setting of four iterations and four subsets was used for all reconstructions [30]. The reconstructed image matrix was 128×128×128 with a voxel size of 3.0 _×_ 3.0 _×_ 3.0 mm^3^. Poisson noise was inherently modeled in the SIMIND simulations and carried out through the reconstruction process. Scatter correction (for SC and ACSC images) was integrated into the CASTOR reconstruction by providing the scaled scatter projection files (generated by SIMIND for each DEW or TEW configuration) as an additive component within CASTOR’s forward projection model during the iterative process. This method incorporates the scatter estimate into the statistical model of the reconstruction rather than performing a simple pre-reconstruction subtraction, which can help preserve statistical properties and potentially reduce noise amplification. For ACSC reconstructions, attenuation correction was performed using the attenuation maps (in units of cm^−1^) generated by the XCAT phantom for each frame, which were also processed by SIMIND and provided to CASTOR. Following reconstruction, images were processed and visualized using AMIDE (A Medical Image Data Examiner) software [31]. Prior studies in myocardial perfusion imaging have shown that Butterworth filters with orders of 3-9 and cutoff frequencies of 0.3-0.9 provide optimal contrast without compromising diagnostic accuracy [32]. To identify the most suitable parameters for our dataset, we systematically tested various filter configurations on simulated images across all frames and correction methods (UN, SC, ACSC), including all scatter correction techniques. Based on this evaluation, a Butterworth filter with an order of 3 and a cutoff frequency of 0.5 achieved the best balance between noise reduction and spatial resolution, maximizing defect contrast, signal-to-noise ratio (SNR) and contrast-to-noise ratio (CNR). These settings were applied to experimental data later on.

### 2.6 Image Quality Assessment

Quantitative assessment of image quality was performed on the reconstructed datasets for all eight frames and all correction conditions. Three distinct Regions of Interest (ROIs) were defined for this analysis: the LV myocardium (healthy), representing normal, well-perfused myocardial tissue; the anterolateral lesion (defect), corresponding to the hypo-perfused region; and the background, selected from a region outside the heart to assess image noise. To ensure consistency and minimize operator variability, ROIs for the normal myocardium and the anterolateral lesion were defined anatomically based on the XCAT phantom’s mid-slice short-axis view. These template ROIs were then precisely overlaid onto the corresponding reconstructed images for each frame. The background ROI was defined manually in a consistent location in each reconstructed image to accurately capture noise characteristics in an area without significant tracer uptake. Representative ROI placements are shown in Figure 2. The following image quality metrics were evaluated to assess the performance of correction techniques.

**Figure 2:**
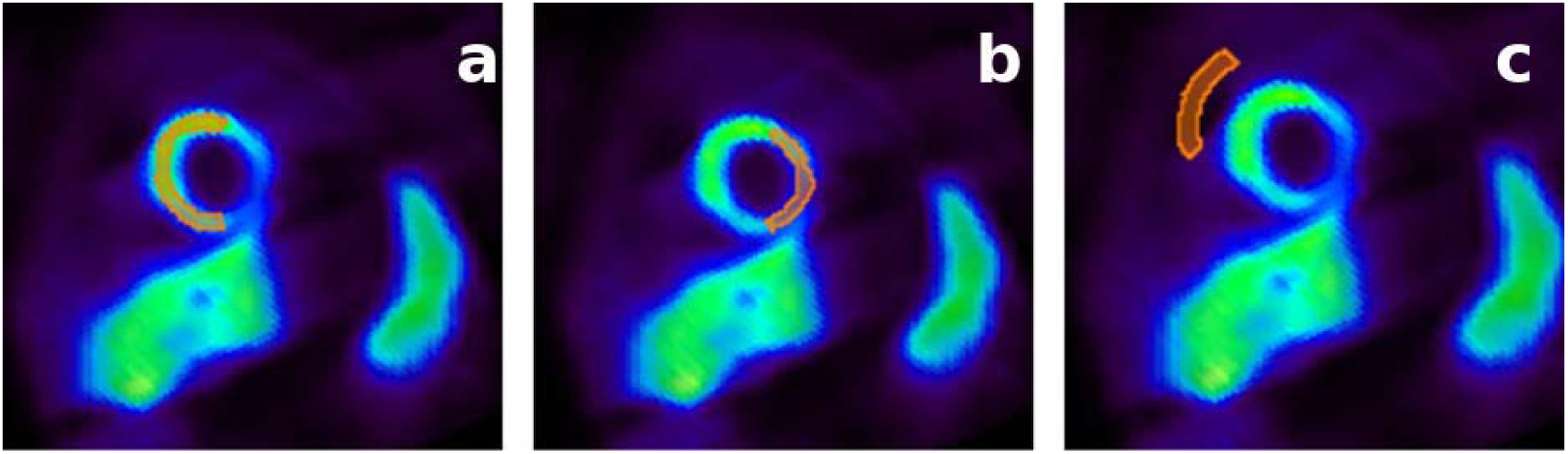
Representative Regions of Interest (ROIs) defined for image quality assessment, shown on a short-axis view of a reconstructed myocardial perfusion SPECT image: (a) ROI encompassing healthy myocardium, (b) ROI targeting the anterolateral perfusion defect, and (c) ROI in the background region.

**Defect contrast (%)** quantifies the relative difference in tracer uptake between healthy myocardium and the perfusion defect. Calculated as shown in Equation (4) [33], where µ_MY ONorm_ is the mean pixel value in the healthy myocardium ROI and µ_MY ODef_ is the mean pixel value in the defect ROI. A higher positive defect contrast value indicates a greater difference in tracer concentration, suggesting a more pronounced perfusion deficit.

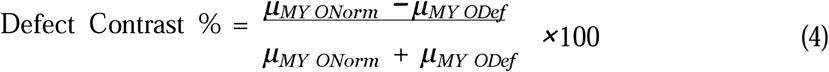

**Contrast-to-Noise Ratio (CNR)** is a widely used metric to quantitatively assess the perfusion defect from its surrounding background (the normal myocardium) in an image. It was calculated using the following Equation (5) [34], where *σ_MY ONorm_* is the standard deviation of pixel values in the healthy myocardium ROI. A higher standard deviation indicates greater fluctuations in pixel values, implying more noise. Therefore, the CNR essentially normalizes the signal difference by the noise level in the normal myocardium. A higher CNR value suggests that the contrast between the defect and the normal myocardium is stronger relative to the noise, making the defect more easily detectable and quantifiable.

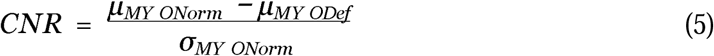

The **signal-to-noise ratio (SNR)** is another key indicator of image quality, used to quantitatively assess the quality an image by comparing the level of the desired signal to the level of background noise. It was calculated using the following Equation 6 [11]. A higher SNR value indicates that the signal from the normal myocardium is significantly stronger than the background noise, resulting in a less noisy image of the healthy myocardium.

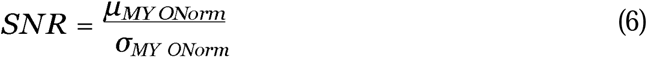

Finally, the **relative noise to background (RNB)** metric assesses how noisy an image is relative to the background activity level. It was calculated using the following Equation 7 [7], where *σ_BG_* is the standard deviation and µ_BG_ is the mean pixel value in the background ROI. Lower RNB values are desirable, indicating a cleaner background. A lower RNB value is generally desirable, as it indicates that the noise level in the background is small relative to the average background signal. This suggests a cleaner background with less random fluctuation, which can improve the overall visual quality of the image and potentially enhance the detectability of features or low-contrast lesions.

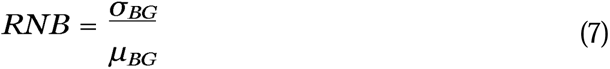

### 2.7 Statistical Analysis

Since the image quality metric data was not normally distributed, non-parametric statistical tests were done. The Friedman test was used for comparing more than two related groups (e.g., UC vs. SC vs. ACSC, or different TEW window widths). When the Friedman test indicated a significant difference, post-hoc pairwise comparisons were conducted using Dunn’s test with Bonferroni correction for multiple comparisons. The Wilcoxon Signed-Rank test was used for comparing two related groups (e.g., TEW overlap vs. non-overlap, DEW SC vs. DEW ACSC, or DEW vs. TEW averaged). A p-value less than 0.05 was considered_√_<u>st</u>atistically significant. Effect sizes r were calculated for Wilcoxon tests where r = Z/ N, where N is the total number of observations (pairs).

## 3 Results

### 3.1 Effect of Correction Method on Image Quality Parameters

The influence of the three primary correction methods: Uncorrected (UC), Scatter Correction only (SC), and combined Attenuation and Scatter Correction (ACSC), on image quality was evaluated across all simulated frames and scatter correction techniques (DEW and all TEW configurations). A Friedman test revealed statistically significant differences among the three correction methods for all four image quality metrics: defect contrast (*χ*^2^ = 220.07, *p* < 0.001), CNR (*χ^2^* = 188.64, *p* < 0.001), SNR (*χ*^2^ = 183.82, *p* < 0.001), and RNB (*χ*^2^ = 171.5, *p* < 0.001). Descriptive statistics including mean, standard deviation (SD), median, and mean ranks for each metric under each correction method are presented in Table 7

**Table 7:** Descriptive Statistics and Mean Ranks for Image Quality Metrics by Correction Method.

| Metric | Group | Mean ± SD | Median | Mean Rank |
| --- | --- | --- | --- | --- |
| Defect Contrast (%) | UC | 22.24 ± 1.53 | 22.71 | 1.98 |
| | SC | 21.53 ± 1.45 | 21.96 | 1.02 |
| | ACSC | 31.18 ± 1.20 | 31.40 | 3.00 |
| CNR | UC | 10.06 ± 2.18 | 10.33 | 1.20 |
| | SC | 11.21 ± 2.93 | 10.65 | 1.80 |
| | ACSC | 22.38 ± 8.52 | 20.77 | 3.00 |
| SNR | UC | 27.45 ± 4.93 | 27.79 | 1.06 |
| | SC | 31.34 ± 6.81 | 29.58 | 2.07 |
| | ACSC | 46.48 ± 16.93 | 40.89 | 2.87 |
| RNB | UC | 0.95 ± 0.07 | 0.95 | 3.00 |
| | SC | 0.34 ± 0.03 | 0.34 | 1.63 |
| | ACSC | 0.34 ± 0.06 | 0.33 | 1.38 |

Post-hoc pairwise comparisons using Dunn’s test with Bonferroni correction (Table 8) further explained these differences. Regarding defect contrast, ACSC significantly improved defect contrast compared to both UC (mean difference +40.2%, *p* < 0.001) and SC (mean difference +44.8 %, *p* < 0.001). SC resulted in a statistically significant, albeit small, decrease in defect contrast compared to UC (mean difference -3.2%, *p* < 0.001). In terms of CNR, ACSC provided significantly higher CNR than UC (mean difference +122.5%, *p* < 0.001) and SC (mean difference +99.7%, *p* < 0.001). SC also showed a significant improvement in CNR over UC (mean difference +11.4%, *p* < 0.001). For SNR, ACSC significantly enhanced SNR compared to UC (mean difference +69.3%, *p* < 0.001) and SC (mean difference +48.3%, *p* < 0.001). SC significantly improved SNR over UC (mean difference +14.2%, *p* < 0.001). Concerning RNB, both SC and ACSC presented a marked decrease of 64.2% relative to UC, with no significant difference observed between SC and ACSC. (*p* = 0.184). These results are visually summarized in Figure 3, which presents boxplots for each image quality metric across the UC, SC, and ACSC correction methods.

**Figure 3:**
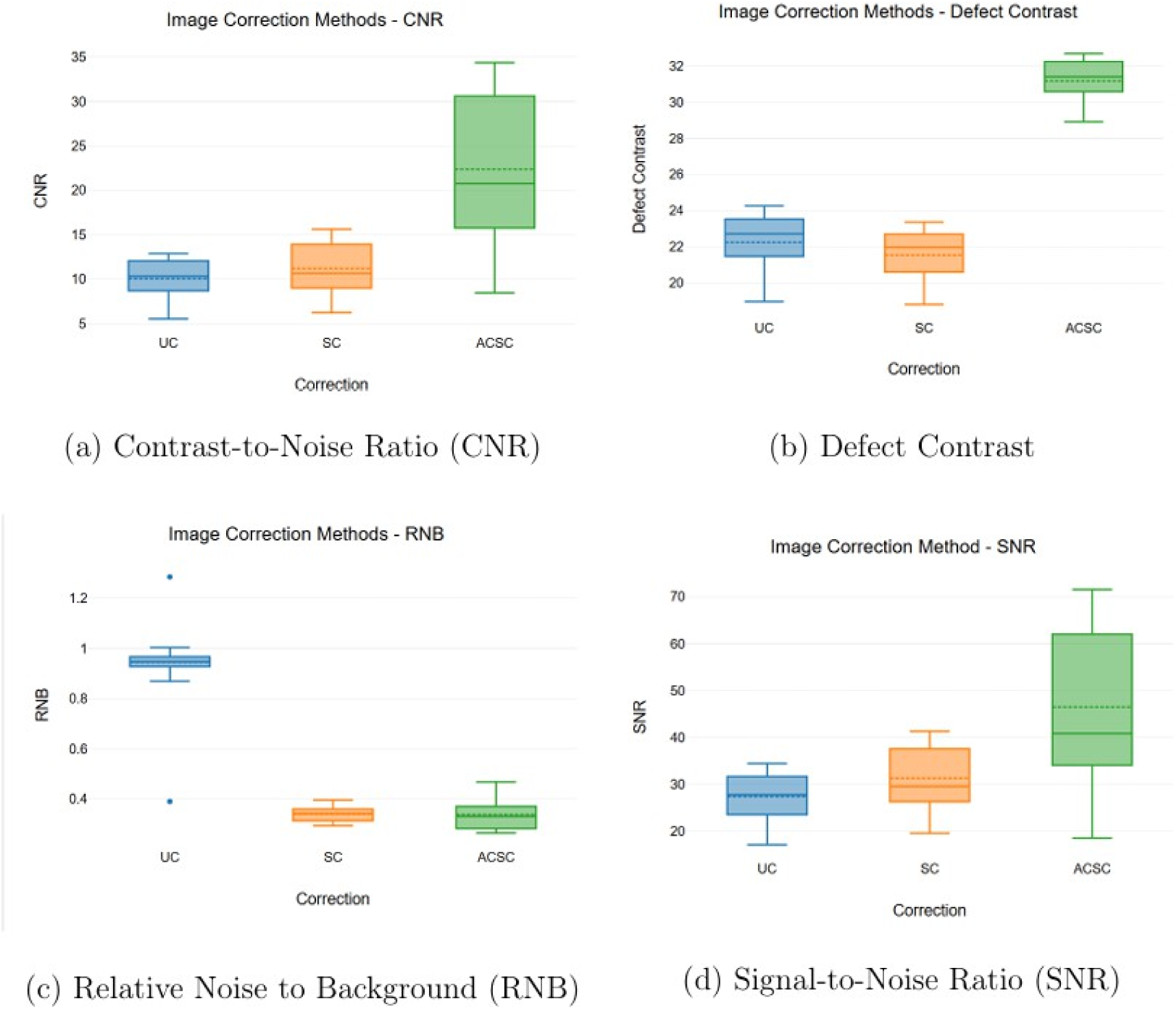
Distribution of image quality metrics across the three correction methods (UC: Uncorrected, SC: Scatter Corrected, ACSC: Attenuation and Scatter Corrected). a) Contrast-to-Noise-Ratio (CNR) b) Defect Contrast c) Relative Noise to Background (RNB) d) Signal-to-Noise Ratio (SNR).

**Table 8:** Dunn-Bonferroni Post-hoc Pairwise Comparisons for Correction Methods.

| Metric | Comparison | Test Statistic (Mean Rank Diff.) | Z-score | Adjusted $p$-value |
| --- | --- | --- | --- | --- |
| Defect Contrast | UC vs SC | 0.96 | 7.22 | < 0.001 |
|  | UC vs ACSC | -1.02 | -7.62 | < 0.001 |
|  | SC vs ACSC | -1.98 | -14.83 | < 0.001 |
| CNR | UC vs SC | -0.61 | -4.54 | < 0.001 |
|  | UC vs ACSC | -1.80 | -13.50 | < 0.001 |
|  | SC vs ACSC | -1.20 | -8.95 | < 0.001 |
| SNR | UC vs SC | -1.00 | -7.52 | < 0.001 |
|  | UC vs ACSC | -1.81 | -13.53 | < 0.001 |
|  | SC vs ACSC | -0.80 | -6.01 | < 0.001 |
| RNB | UC vs SC | 1.38 | 10.29 | < 0.001 |
|  | UC vs ACSC | 1.63 | 12.16 | < 0.001 |
|  | SC vs ACSC | 0.25 | 1.87 | 0.184 |

### 3.2 Effect of TEW Scatter Window Widths

The impact of varying the TEW scatter window widths (2 keV, 4 keV, and 6 keV) was assessed separately for images corrected with SC only and for images corrected with ACSC, combining data from both overlapping and non-overlapping configurations. For SC images (Table 9), the Friedman test indicated no statistically significant differences among the three window widths for defect contrast (χ^2^ = 1.75, p = 0.417), SNR (χ^2^ = 1.75, p = 0.417), or RNB (χ^2^ = 4.19, p = 0.123). For CNR, the result approached significance (χ^2^ = 5.81, p = 0.055) but did not meet the conventional threshold. For ACSC images (Table 10), the Friedman test also showed no statistically significant differences among the window widths for defect contrast (χ^2^ = 2.31, p = 0.315), CNR (χ^2^ = 2.69, p = 0.261), SNR (χ^2^ = 0.75, p = 0.687), or RNB (χ^2^ = 1.75, p = 0.417). Overall, within the tested range, varying the TEW scatter window width from two to six keV did not have a statistically meaningful impact on the evaluated image quality metrics, regardless of whether only scatter correction or combined attenuation and scatter correction was applied.

**Table 9:** Comparison of Image Quality Metrics Across TEW Window Widths for SC Images.

| Metric | Window Width | Mean ± SD | Median | Mean Rank | $\chi^2$ | p-value |
| --- | --- | --- | --- | --- | --- | --- |
| Defect Contrast (%) | 2 keV | 21.53 ± 1.46 | 22.00 | 1.94 | 1.75 | 0.417 |
| | 4 keV | 21.52 ± 1.49 | 21.95 | 1.88 | | |
| | 6 keV | 21.53 ± 1.46 | 21.99 | 2.19 | | |
| CNR | 2 keV | 11.17 ± 2.95 | 10.63 | 1.88 | 5.81 | 0.055 |
| | 4 keV | 11.14 ± 2.94 | 10.76 | 1.78 | | |
| | 6 keV | 11.30 ± 3.00 | 11.06 | 2.34 | | |
| SNR | 2 keV | 31.22 ± 6.89 | 29.43 | 1.94 | 1.75 | 0.417 |
| | 4 keV | 31.16 ± 6.82 | 29.80 | 1.88 | | |
| | 6 keV | 31.58 ± 6.95 | 31.01 | 2.19 | | |
| RNB | 2 keV | 0.34 ± 0.03 | 0.34 | 2.22 | 4.19 | 0.123 |
| | 4 keV | 0.34 ± 0.03 | 0.35 | 2.06 | | |
| | 6 keV | 0.34 ± 0.03 | 0.34 | 1.72 | | |

**Table 10:** Comparison of Image Quality Metrics Across TEW Window Widths for ACSC Images.

| Metric | Window Width | Mean ± SD | Median | Mean Rank | $\chi^2$ | p-value |
| --- | --- | --- | --- | --- | --- | --- |
| Defect Contrast (%) | 2 keV | 31.17 ± 1.21 | 31.43 | 1.91 | 2.31 | 0.315 |
| | 4 keV | 31.18 ± 1.25 | 31.36 | 1.88 | | |
| | 6 keV | 31.19 ± 1.21 | 31.45 | 2.22 | | |
| CNR | 2 keV | 22.26 ± 8.58 | 20.67 | 1.78 | 2.69 | 0.261 |
| | 4 keV | 22.38 ± 8.61 | 21.08 | 2.19 | | |
| | 6 keV | 22.53 ± 8.71 | 21.66 | 2.03 | | |
| SNR | 2 keV | 46.49 ± 17.17 | 42.73 | 1.88 | 0.75 | 0.687 |
| | 4 keV | 45.89 ± 16.87 | 40.69 | 2.06 | | |
| | 6 keV | 47.00 ± 17.35 | 45.06 | 2.06 | | |
| RNB | 2 keV | 0.34 ± 0.06 | 0.34 | 2.06 | 1.75 | 0.417 |
| | 4 keV | 0.34 ± 0.06 | 0.33 | 1.81 | | |
| | 6 keV | 0.34 ± 0.06 | 0.32 | 2.13 | | |

### 3.3 Effect of Overlapping vs. Non-Overlapping TEW Scatter Windows

The Wilcoxon Signed-Rank test was used to compare TEW performance with overlapping versus non-overlapping scatter windows. This comparison pooled data across all three window widths (2, 4, and 6 keV) and all eight simulated frames (each run twice). For SC images (Table 11), no statistically significant differences were observed between overlapping and non-overlapping conditions for defect contrast (*W* = 549, *Z* =_−_0.4, p = 0.689, *r* = 0.06), CNR (*W* = 588, Z = 0.0, *p* = 1.000, *r* = 0.00), SNR (*W* = 567, *Z* = _−_ 0.22, *p* = 0.829, *r* = 0.03), or RNB (*W* = 534, *Z* = _−_0.55, *p* = 0.580, *r* = 0.08). Similarly, for ACSC images (Table 12), no statistically significant differences were found for defect contrast (*W* = 500, *Z* = −0.68, *p* = 0.498, *r* = 0.10), CNR (*W* = 542, Z = −0.23, *p* = 0.816, *r* = 0.03), SNR (*W* = 558, *Z* = −0.06, *p* = 0.949, *r* = 0.01), or RNB (*W* = 501, *Z* = −0.67, *p* = 0.505, *r* = 0.10). In all cases, the effect sizes (r) were very small, indicating negligible practical differences between using overlapping or non-overlapping TEW scatter windows for the configurations tested.

**Table 11:** Comparison Between SC Overlap and Non-Overlap TEW Conditions.

| Metric | Group | Mean ±SD | Median | W | Z-score | p-value | r |
| --- | --- | --- | --- | --- | --- | --- | --- |
| Defect Contrast (%) | Overlap | 21.54 ± 1.44 | 21.95 | 549 | -0.40 | 0.689 | 0.06 |
| | Non-Overlap | 21.51 ± 1.48 | 21.95 | | | | |
| CNR | Overlap | 11.19 ± 2.90 | 10.65 | 588 | 0.00 | 1.000 | 0.00 |
| | Non-Overlap | 11.22 ± 3.00 | 10.85 | | | | |
| SNR | Overlap | 31.28 ± 6.73 | 29.53 | 567 | -0.22 | 0.829 | 0.03 |
| | Non-Overlap | 31.36 ± 6.97 | 30.03 | | | | |
| RNB | Overlap | 0.34 ± 0.03 | 0.34 | 534 | -0.55 | 0.580 | 0.08 |
| | Non-Overlap | 0.34 ± 0.03 | 0.34 | | | | |

**Table 12:** Comparison Between ACSC Overlap and Non-Overlap TEW Conditions.

| Metric | Group | Mean ±SD | Median | W | Z-score | p-value | r |
| --- | --- | --- | --- | --- | --- | --- | --- |
| Defect Contrast (%) | Overlap | 31.20 ± 1.20 | 31.40 | 500 | -0.68 | 0.498 | 0.10 |
| | Non-Overlap | 31.16 ± 1.24 | 31.40 | | | | |
| CNR | Overlap | 22.39 ± 8.49 | 20.77 | 542 | -0.23 | 0.816 | 0.03 |
| | Non-Overlap | 22.39 ± 8.68 | 20.95 | | | | |
| SNR | Overlap | 46.16 ± 16.75 | 40.89 | 558 | -0.06 | 0.949 | 0.01 |
| | Non-Overlap | 46.76 ± 17.33 | 43.36 | | | | |
| RNB | Overlap | 0.34 ± 0.06 | 0.33 | 501 | -0.67 | 0.505 | 0.10 |
| | Non-Overlap | 0.34 ± 0.06 | 0.33 | | | | |

### 3.4 DEW Method: Scatter Correction (SC) versus Attenuation and Scatter Correction (ACSC)

The impact of adding attenuation correction to DEW scatter correction was evaluated using the Wilcoxon Signed-Rank test across all eight frames (each simulated twice). As summarized in Table 13 and illustrated in Figure 4, DEW ACSC resulted in significantly higher defect contrast (*W* = 0, *Z* = –3.52, *p* < 0.001, *r* = 0.88), CNR (*W* = 0, *Z* = – 3.52, *p* < 0.001, *r* = 0.88), and SNR (*W* = 3, *Z* = –3.36, *p* = 0.001, *r* = 0.84) compared to DEW SC alone. The effect sizes were large, indicating significant improvements with the addition of AC. No statistically significant difference was observed for RNB (*W* = 53, *Z* = –0.78, *p* = 0.438, *r* = 0.19), suggesting RNB was primarily influenced by scatter correction itself.

**Figure 4:**
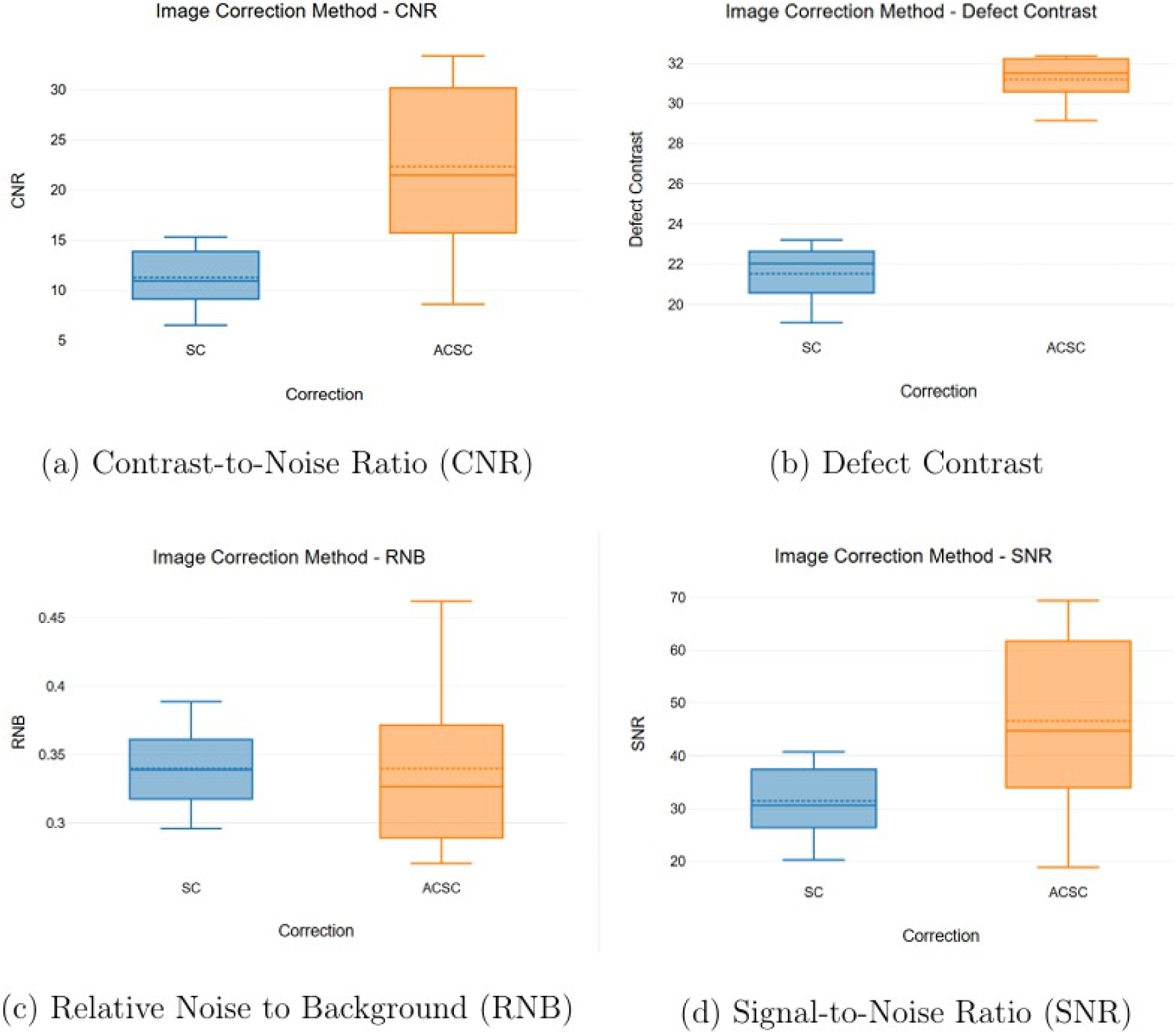
Comparison of image quality metrics for the Dual Energy Window (DEW) method with Scatter Correction only (SC) versus combined Attenuation and Scatter Correction (ACSC).

**Table 13:** Comparison of DEW SC vs. DEW ACSC for Image Quality Metrics.

| Metric | Group | Mean ±SD | Median | W | Z-score | p-value | r |
| --- | --- | --- | --- | --- | --- | --- | --- |
| Defect Contrast (%) | SC | 21.53 ± 1.44 | 22.04 | 0 | -3.52 | < 0.001 | 0.88 |
| | ACSC | 31.19 ± 1.17 | 31.51 | | | | |
| CNR | SC | 11.25 ± 3.01 | 10.91 | 0 | -3.52 | < 0.001 | 0.88 |
| | ACSC | 22.34 ± 8.69 | 21.48 | | | | |
| SNR | SC | 31.45 ± 6.98 | 30.59 | 3 | -3.36 | 0.001 | 0.84 |
| | ACSC | 46.61 ± 17.32 | 44.75 | | | | |
| RNB | SC | 0.34 ± 0.03 | 0.34 | 53 | -0.78 | 0.438 | 0.19 |
| | ACSC | 0.34 ± 0.06 | 0.33 | | | | |

### 3.5 Comparison of TEW and DEW Scatter Correction Methods

To compare the overall performance of TEW and DEW methods, TEW results were averaged across all its window width and overlap configurations, justified by the lack of significant differences within these TEW subgroups (subsections 3.2 and 3.3). Wilcoxon Signed-Rank tests were then performed. Under the SC only condition (Table 14), no statistically significant differences were observed between the averaged TEW (SC-avg TEW) and DEW (SC-DEW) methods for defect contrast (*W* = 59, *Z* = –0.47, *p* = 0.642, *r* = 0.12), CNR (*W* = 57, *Z* = –0.57, *p* = 0.569, *r* = 0.14), SNR (*W* = 55, *Z* = –0.67, *p* = 0.501, *r* = 0.17), or RNB (*W* = 61, *Z* = –0.36, *p* = 0.717, *r* = 0.09). Similarly, under the ACSC condition (Table 15), no statistically significant differences were found between averaged TEW (ACSC-avg TEW) and DEW (ACSC-DEW) for defect contrast (*W* = 62, *Z* = –0.31, *p* = 0.756, *r* = 0.08), CNR (*W* = 56, *Z* = –0.62, *p* = 0.535, *r* = 0.16), SNR (*W* = 67, *Z* = –0.05, *p* = 0.959, *r* = 0.01), or RNB (*W* = 46, *Z* = –1.14, *p* = 0.255, *r* = 0.28). The effect sizes in all comparisons were small, suggesting that, on average, DEW and TEW provide comparable image quality across the metrics evaluated, regardless of whether attenuation correction is concurrently applied. Representative reconstructed images for vertical long axis (VLA), horizontal long axis (HLA), and short axis (SA) views across all correction methods and frames are shown in Figure 5.

**Figure 5:**
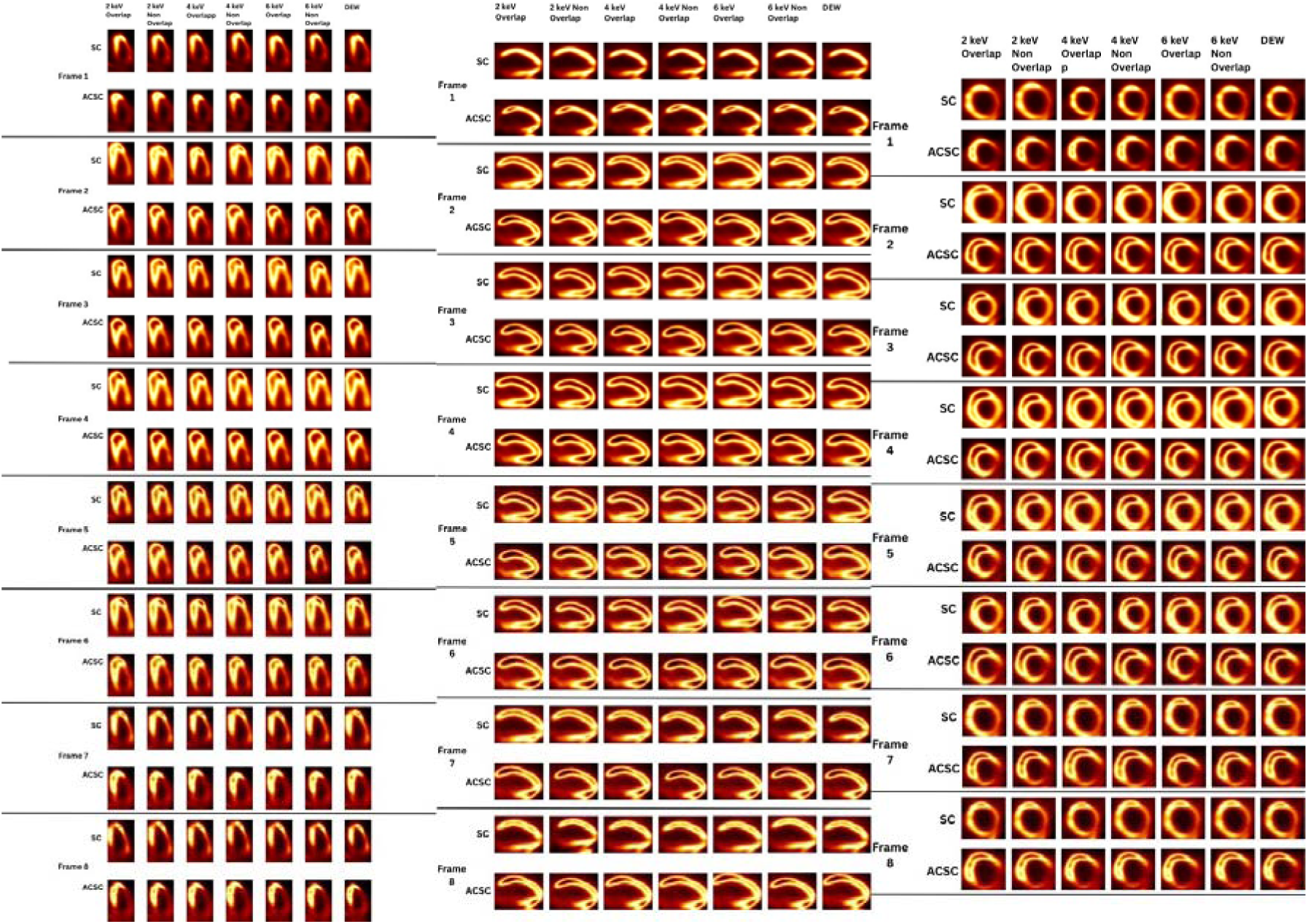
Vertical long axis (VLA), horizontal long axis (HLA), and short axis (SA) views of reconstructed images for all 8 frames across different correction methods (SC, ACSC) and energy window configurations.

**Table 14:** Comparison Between Averaged TEW (SC) and DEW (SC) Groups.

| Metric | Group | Mean ±SD | Median | W | Z-score | p-value | r |
| --- | --- | --- | --- | --- | --- | --- | --- |
| Defect Contrast (%) | SC-avg TEW | 21.53 ± 1.49 | 21.99 | 59 | -0.47 | 0.642 | 0.12 |
| | SC-DEW | 21.53 ± 1.44 | 22.04 | | | | |
| CNR | SC-avg TEW | 11.20 ± 3.00 | 10.97 | 57 | -0.57 | 0.569 | 0.14 |
| | SC-DEW | 11.25 ± 3.01 | 10.91 | | | | |
| SNR | SC-avg TEW | 31.32 ± 6.97 | 30.45 | 55 | -0.67 | 0.501 | 0.17 |
| | SC-DEW | 31.45 ± 6.98 | 30.59 | | | | |
| RNB | SC-avg TEW | 0.34 ± 0.03 | 0.34 | 61 | -0.36 | 0.717 | 0.09 |
| | SC-DEW | 0.34 ± 0.03 | 0.34 | | | | |

**Table 15:** Comparison Between Averaged TEW (ACSC) and DEW (ACSC) Groups.

| Metric | Group | Mean ±SD | Median | W | Z-score | p-value | r |
| --- | --- | --- | --- | --- | --- | --- | --- |
| Defect Contrast (%) | ACSC-avg TEW | 31.18 ± 1.24 | 31.42 | 62 | -0.31 | 0.756 | 0.08 |
| | ACSC-DEW | 31.19 ± 1.17 | 31.51 | | | | |
| CNR | ACSC-avg TEW | 22.39 ± 8.76 | 21.28 | 56 | -0.62 | 0.535 | 0.16 |
| | ACSC-DEW | 22.34 ± 8.69 | 21.48 | | | | |
| SNR | ACSC-avg TEW | 46.46 ± 17.19 | 44.09 | 67 | -0.05 | 0.959 | 0.01 |
| | ACSC-DEW | 46.61 ± 17.32 | 44.75 | | | | |
| RNB | ACSC-avg TEW | 0.34 ± 0.06 | 0.33 | 46 | -1.14 | 0.255 | 0.28 |
| | ACSC-DEW | 0.34 ± 0.06 | 0.33 | | | | |

## 4. Discussion

### 4.1 Key Findings

Our Monte Carlo simulation study reinforces the crucial role of advanced correction techniques in enhancing the quantitative accuracy and overall quality of myocardial perfusion SPECT (MPI).

Foremost among our observations is the consistent and statistically significant superiority of combined attenuation and scatter correction (ACSC) across all evaluated image quality metrics: defect contrast, contrast-to-noise ratio (CNR), signal-to-noise ratio (SNR), and relative noise to background (RNB) (all p < 0.001 when compared to uncorrected or SC-only images). This underscores the interactive benefit of addressing both major photon-degrading phenomena. Specifically, ACSC led to a considerable 122.5% improvement in CNR and a 69.3% increase in SNR relative to uncorrected (UC) images, indicating significantly enhanced lesion detectability against background noise and improved signal consistency within the healthy myocardium. Furthermore, defect contrast improved by 40.2% compared to UC and by a notable 44.8% compared to SC alone, highlighting that attenuation correction is crucial not just for quantitative accuracy but also for optimizing the visual conspicuity of perfusion defects.

The application of scatter correction alone (SC) offered considerable benefits over uncorrected images in terms of noise-related metrics. However, a crucial observation was the slight but statistically significant decrease (approximately 3.2%) in defect contrast when SC was applied without attenuation correction (AC). SC alone led to a 64.2% reduction in RNB, and increases of 14.2% in SNR and 11.4% in CNR. This reflects a classic noise–contrast trade-off in nuclear imaging, where improvements in background uniformity can inadvertently suppress diagnostically relevant signal differences. In this study, scatter was estimated using the Dual Energy Window (DEW) method with a fixed scaling factor (k = 0.5), and this estimate was incorporated as an additive component in the forward model during OSEM iterative reconstruction. This method preserves the Poisson statistics of the measured data and avoids the noise amplification typically associated with projection-based subtraction. However, in the absence of AC, the forward model lacks depth-dependent compensation for photon attenuation, potentially leading to an imbalance between the estimated primary and scatter components. This effect is particularly pronounced in regions of high attenuation or low tracer uptake, such as the inferior myocardial wall, where iterative updates may underestimate activity, flattening the intensity gradient between ischemic and healthy tissue and ultimately reducing defect contrast. Simultaneously, the increases in CNR and SNR are partly explained by reduced variability (standard deviation) in the healthy myocardium region, which lowers the denominator in both metrics. Thus, even if the actual signal difference is smaller, the improved noise characteristics inflate these ratios. Similar patterns were reported by Hosny et al. (2020) [10], where SC-alone reconstructions showed higher noise-based metrics but diminished visual contrast. Furthermore, Noori-Asl et al. (2014) [7] demonstrated that projection-based TEW correction—despite improving contrast—resulted in decreased SNR and increased RNB, further highlighting the risk of image quality trade-offs when scatter correction is applied without concurrent attenuation modeling. These findings collectively emphasize that even when scatter correction is implemented within a statistically integrated reconstruction framework, its isolated application may introduce quantitative biases, reinforcing the necessity of combined attenuation and scatter correction (ACSC) to ensure optimal lesion detection and accurate quantification in myocardial perfusion SPECT imaging.

To address the observed reduction in defect contrast with SC alone, strategies can be considered to refine both scatter estimation and reconstruction modeling. First, using a fixed scaling factor (k = 0.5) in the DEW method may not adequately account for regional differences in scatter behavior across heterogeneous anatomical structures. Implementing a spatially adaptive scaling approach, where k varies based on local activity or anatomical priors, could reduce the risk of overestimating scatter in highly attenuated regions. Additionally, optimizing the configuration of the energy windows used for scatter estimation (e.g., width, position, and overlap with the photopeak window) based on phantom calibration or simulation-based sensitivity analysis could help preserve contrast. Beyond scatter estimation, reconstruction-level strategies can also mitigate contrast loss. One effective approach is the integration of resolution recovery (RR) into the iterative reconstruction process. By modeling the system’s point spread function, RR can enhance spatial resolution and restore edge sharpness, which is especially beneficial for improving the visibility of perfusion defects that may appear less distinct in SC-only images. Another important consideration is the optimization of OSEM reconstruction parameters—particularly the number of iterations and subsets. Excessive iterations in the SC-only condition may amplify modeling inconsistencies or suppress true signal differences, leading to further contrast degradation. Adjusting these parameters to balance noise suppression with contrast preservation can help maintain diagnostic image quality. Together, these strategies offer practical means to enhance contrast recovery in SC-only images without introducing additional correction models.

One of the surprising outcomes of this study was the lack of statistically significant impact of varying TEW scatter window widths (from 2 keV to 6 keV) or toggling between overlapped and non-overlapping TEW configurations on any of the image quality metrics. This was true for both SC-only and ACSC images. It is possible that the differences induced by these subtle window geometry changes are minor compared to the dominant effect of applying the scatter correction algorithm itself, or the even larger effect of including attenuation correction. The fixed scaling parameters within SIMIND for the trapezoidal TEW approximation might also contribute to this observed insensitivity. This suggests that for Tc 99m MPI, once the TEW method is applied, fine-tuning these specific window parameters within the 2 keV to 6 keV range may offer diminishing returns in terms of the image quality metrics evaluated here.

Equally noteworthy was the comparison between the DEW (with k = 0.5) and the TEW methods (averaged across all its configurations). No statistically significant differences were observed for any image quality metric, regardless of whether SC was applied alone or in combination with AC. The similar performance across methods also implies that the accuracy of the scatter estimate, once a certain threshold is met, might be less critical than the robust application of AC.

The non-significant difference in RNB between SC and ACSC conditions, despite ACSC showing marked improvements in other metrics, is also an interesting point. It suggests that the primary reduction in relative background noise is achieved by addressing scattered photons. While attenuation correction is vital for restoring quantitative accuracy and improving contrast and SNR by compensating for photon loss, it may not further refine the noise characteristics of the background beyond what scatter correction already achieves.

### 4.2 Clinical Significance

The consistent and statistically significant superiority of combined attenuation and scatter correction (ACSC) demonstrated in this study carries direct and critical implications for clinical practice. The substantial improvements observed with ACSC compared to uncorrected images translate directly to enhanced confidence in lesion detectability and more reliable quantification of myocardial perfusion. Such accuracy is paramount for precise diagnosis, effective risk stratification, and the formulation of optimal therapeutic strategies for patients with suspected or known coronary artery disease. Furthermore, the optimization of defect contrast with ACSC is vital for the clear visual delineation of perfusion abnormalities, ensuring that even subtle defects are not overlooked.

While the application of scatter correction alone (SC) does offer benefits in terms of noise reduction, our findings highlight a critical trade-off: a slight but statistically significant decrease in defect contrast when attenuation correction (AC) is omitted. Clinically, this implies that relying solely on SC might inadvertently mask the true extent or severity of a perfusion defect. This occurs because, without AC, the reconstruction model may inadequately compensate for photon attenuation, potentially leading to an underestimation of activity in certain myocardial regions and a flattening of diagnostically important intensity gradients. Therefore, for achieving optimal diagnostic accuracy and quantitative integrity in myocardial perfusion SPECT, the integration of robust AC with SC is not merely beneficial but essential.

The observation that there were no statistically significant differences between the DEW and TEW scatter correction methods, nor from minor variations in TEW window parameters (within the 2 keV to 6 keV range for Tc 99m MPI), offers valuable practical guidance. It suggests a degree of flexibility in clinical settings: a well-optimized DEW method, often simpler to implement, can provide comparable image quality to the more complex TEW method when both are coupled with AC. This is particularly relevant for clinics aiming to standardize protocols or where resources for intricate TEW parameter optimization are limited. The core clinical takeaway is that the profound benefit arises from the diligent application of both attenuation and a robust scatter correction, rather than from subtle adjustments to the scatter correction technique itself. The primary role of SC is to reduce background noise, while AC is indispensable for restoring quantitative accuracy and maximizing defect conspicuity and signal strength.

### 4.3 Comparison to Literature

The findings of this study align well with prior research advocating for integrated correction strategies to achieve optimal image quality in SPECT MPI. The superiority of ACSC observed here is consistent with studies by Ye et al. (1992) [12] and Hosny et al. (2020) [10], who also demonstrated the benefits of combined corrections.

Our results regarding the improvements in SNR, CNR, and RNB with scatter correction alone diverge somewhat from Noori-Asl et al. (2013) [6] and Noori-Asl et al. (2014) [7], who reported that TEW increased background noise and reduced SNR, or led to a decline in SNR and an increase in RNB despite contrast improvement. Rafati et al. (2017) [8] also indicated superior gains in both contrast and SNR with TEW. This discrepancy may be explained by differences in reconstruction strategy; while the aforementioned studies primarily relied on projection subtraction and FBP-based reconstruction, our approach integrated scatter estimates directly into the forward model of OSEM. This likely mitigated the noise amplification typically observed in conventional correction workflows.

Our observation that SC alone improved noise-related metrics (SNR, CNR, RNB) but led to a slight reduction in defect contrast compared to uncorrected images is consistent with the findings of Hosny et al. (2020) [10]. Both studies emphasize that SC is most effective when integrated with attenuation correction, supporting the superiority of the combined ACSC approach for optimizing all image quality metrics.

Regarding the comparison of TEW window parameters, while some literature, such as Bouzekraoui et al. (2019) [20] and Asmi et al. (2020) [19], has suggested that optimal sub-window settings can be isotope- or setup-dependent, our findings for Tc 99m within the tested range (2 keV to 6 keV) indicate robustness. This suggests that for Tc 99m MPI, the specific TEW window width or overlap configuration (within the tested ranges) may be less critical than the application of the TEW method itself in conjunction with AC.

Alaei et al. (2023) [14], using a similar XCAT and OSEM-based simulation, found that AC + TEW yielded the highest contrast, while AC + DEW achieved the best SNR. In contrast, our results showed no significant difference between DEW and TEW under ACSC conditions, suggesting comparable performance across metrics when both methods are properly integrated. This difference may stem from variations in how TEW and DEW were parameterized (e.g., sub-window widths or energy window placements, which were not explicitly mentioned in their work), or from differences in statistical testing and metric definitions. The comparable performance found between DEW (with k = 0.5) and the averaged TEW methods contributes to a growing body of evidence (e.g., Noori-Asl and Eghbal, 2024 [15]) suggesting that a well-configured DEW can perform similarly to TEW. This challenges the long-standing assumption of TEW’s categorical superiority and suggests that DEW might be an equally practical, simpler option in many clinical scenarios, provided it is properly optimized and, crucially, paired with attenuation correction.

Nonetheless, the broader literature still shows no consensus on the overall consequences of scatter correction and its optimal settings. For instance, Jeong et al. (2001) [16] reported that while DEW improved contrast, it had no significant impact on left ventricular functional measurements. Galt et al. (1992) [17] indicated improved quantification with SC, whereas Khalil et al. (2004) [18] observed only enhanced image contrast with no significant SNR improvements. These varying outcomes underscore the value of our systematic investigation within a highly realistic simulation environment.

### 4.4. Limitations and Future Work

This study presents a detailed evaluation of scatter correction methods using Monte Carlo simulations; however, several limitations should be considered. The analysis was based solely on simulations using the 4D XCAT phantom representing a standard adult male. Although this phantom provides realistic anatomical and physiological features, it does not fully capture the biological variability encountered in clinical settings. Factors such as gender, body mass index, organ size, and the presence of cardiac pathologies can influence attenuation and scatter differently. Future work should incorporate phantoms with varied anatomies or use patient-specific simulations to improve the generalizability of the findings.

Another limitation is the use of fixed parameters in both correction and reconstruction steps. The Dual Energy Window (DEW) method was implemented with a constant scaling factor (k = 0.5), commonly reported in literature, yet this value may not be optimal for all imaging conditions. Similarly, the Triple Energy Window (TEW) technique relied on the built-in trapezoidal approximation in SIMIND, and other TEW approximations such as the triangular one were not explored. Investigating adaptive or data-driven approaches for determining the scaling factor and testing alternative TEW configurations could lead to more effective correction strategies.

All reconstructions were performed using a fixed OSEM setup with four iterations, four subsets, and a Butterworth post-filter. While this reflects common practice, the study did not examine how variations in reconstruction parameters or algorithms affect image quality. In particular, resolution recovery and other reconstruction methods, such as filtered back projection (FBP), were not included. A comparative analysis of different reconstruction techniques and parameter settings would help clarify how reconstruction interacts with scatter and attenuation correction.

In terms of energy window selection, the TEW method was tested with three discrete window widths (2, 4, and 6 keV). Although these represent typical values, wider sub-windows or asymmetric sampling of window configurations may uncover different patterns not observed in this study.

Finally, the evaluation of image quality focused exclusively on quantitative metrics such as defect contrast, contrast-to-noise ratio (CNR), signal-to-noise ratio (SNR), and relative noise to background (RNB). While these metrics are informative, they do not fully capture clinical relevance. Future studies should incorporate diagnostic task-based evaluations, including lesion detectability studies, human or model observer assessments, and receiver operating characteristic (ROC) analysis, to better connect quantitative image quality with diagnostic performance.

## 5. Conclusion

This Monte Carlo simulation study systematically evaluated the effects of DEW and TEW scatter correction techniques, in conjunction with attenuation correction, on quantitative Tc 99m myocardial perfusion SPECT imaging. The results clearly demonstrate that a comprehensive correction strategy combining both attenuation and scatter correction (ACSC) yields the best image quality, significantly enhancing defect contrast, CNR, and SNR compared to uncorrected images or those with scatter correction alone.

A particularly novel finding of this study is the identification of a trade-off with SC alone: although it improved SNR, CNR, and RNB, it led to a statistically significant reduction in defect contrast. This effect—previously underemphasized in myocardial perfusion SPECT literature, highlights the risk of over-smoothing diagnostically important features when attenuation correction is omitted. This observation, supported by detailed simulation and statistical analysis, suggests that protocols using SC alone should be interpreted with caution, and future studies should investigate adaptive strategies to mitigate contrast loss under such conditions.

This work addresses several unresolved issues in myocardial perfusion imaging. Prior studies have reported conflicting conclusions regarding the relative performance of DEW and TEW, with some favoring one method over the other depending on imaging conditions. However, the effects of window width and overlap configuration in Tc 99m cardiac imaging have not been directly examined. This study fills that gap by exploring these factors using realistic cardiac phantom simulations and provides new insights to support more effective scatter correction practices in myocardial perfusion SPECT. To our knowledge, this is the first study to evaluate the combined influence of window width and overlap geometry on image quality in Tc 99m MPI using advanced Monte Carlo simulations with a realistic 4D XCAT phantom—contributing novel insights to the ongoing debate in the literature.

In addition, the interplay between scatter and attenuation correction under iterative reconstruction algorithms had not been clearly delineated. By examining these factors together using OSEM-based reconstruction and a comprehensive set of image quality metrics, this study helps clarify their individual and combined contributions to overall image performance.

No statistically significant differences were observed between DEW and TEW methods, nor did variations in TEW window width or overlap configuration meaningfully affect the evaluated metrics. These findings suggest that, once optimized, the choice of scatter correction method is less critical than ensuring attenuation correction is incorporated—a key factor for restoring quantitative accuracy and improving lesion detectability. The absence of measurable impact from TEW overlap configurations also has practical implications, supporting a more streamlined protocol design by reducing complexity without compromising diagnostic quality.

In summary, this study advances both theoretical and applied understanding of correction strategies in cardiac SPECT imaging. It shows that a well-implemented DEW method, when combined with AC, performs comparably to TEW, offering a simpler and effective alternative. More importantly, it emphasizes that future protocol development should focus on optimizing correction combinations and reconstruction models, rather than narrowly adjusting scatter correction parameters. These findings support the creation of standardized, evidence-based imaging protocols and promotes more informed clinical and research decision-making in SPECT MPI.

## Data Availability

The data are based on simulation obtained by the student Mounia El Bab

## Author Contributions

Conceptualization, M.E.B. and A.G.; methodology, M.E.B. and A.G.; software, M.E.B.; validation, M.E.B. and A.G.; formal analysis, M.E.B. and A.G.; investigation, M.E.B.; resources, A.G.; data curation, M.E.B.; writing—original draft preparation, M.E.B.; writing—review and editing, M.E.B. and A.G.; visualization, M.E.B.; supervision, A.G.; project administration, A.G. All authors have read and agreed to the published version of the manuscript.

## Abbreviations

AC: Attenuation Correction.
ACSC: Attenuation and Scatter Corrected.
BMI: Body Mass Index.
CAD: Coronary Artery Disease.
CNR: Contrast-to-Noise Ratio.
CO: Cardiac Output.
DEW: Dual Energy Window.
EDV: End-Diastolic Volume.
ESV: End-Systolic Volume.
FBP: Filtered Back Projection.
LEHR: Low Energy High Resolution.
LPO: Left Posterior Oblique.
LV: Left Ventricle / Left Ventricular.
MBq: Megabecquerel.
MC: Monte Carlo.
MPI: Myocardial Perfusion Imaging.
NaI(Tl): Sodium Iodide doped with Thallium.
NCAT: NURBS-Based Cardiac-Torso (phantom).
OSEM: Ordered Subsets Expectation Maximization.
RAO: Right Anterior Oblique.
RNB: Relative Noise to Background.
ROI: Region of Interest.
RR: Resolution Recovery.
SC: Scatter Corrected.
SIMIND: Simulating Medical Imaging Nuclear Detectors (MC Software).
SNR: Signal-to-Noise Ratio.
SPECT: Single Photon Emission Computed Tomography.
SV: Stroke Volume.
TEW: Triple Energy Window.
UC: Uncorrected.
XCAT: Extended Cardiac-Torso (phantom).

## Acknowledgements

This study was made possible through the use of the XCAT phantom program, generously made available by Professor Paul Segars (Duke University), and the SIMIND Monte Carlo simulation software, with technical insights provided by Dr. Michael Ljungberg. Their contributions significantly supported the implementation and accuracy of the simulation environment used in this research.

